# CEREBRAL MICROBLEEDS ARE ASSOCIATED WITH DEFICITS IN COGNITIVE PROCESSING SPEED AND EXECUTIVE FUNCTIONS IN MIDDLE-AGED ADULTS WITH TYPE 1 DIABETES

**DOI:** 10.1101/2025.06.30.25330540

**Authors:** Iiris Kyläheiko, Aleksi Tarkkonen, Linda Kuusela, Juha Martola, Teemu I. Paajanen, Jussi Virkkala, Per-Henrik Groop, Lena M. Thorn, Turgut Tatlisumak, Jukka Putaala, Daniel Gordin, Hanna Jokinen, the FinnDiane Study Group

**Author notes:** Address for correspondence: Hanna Jokinen, Division of Neuropsychology, Neurocenter, Helsinki University Hospital, PO Box 302, 00029 HUS, Finland. Equal contribution (Daniel Gordin, Hanna Jokinen).

## Abstract

**Background:** Adults with type 1 diabetes (T1D) have an increased risk of developing cerebral small vessel disease (cSVD)-related brain changes already in midlife, yet their significance for cognitive functions remains poorly understood. We investigated the associations between cerebral microbleeds (CMBs), white matter hyperintensities (WMHs), and cognitive functions, including processing speed, executive functions, and episodic memory, in individuals without detected neurological symptoms.

**Methods:** Adults with T1D (n=167; age 46.4±7.7 years) underwent clinical and biochemical evaluations, brain magnetic resonance imaging (MRI), and a comprehensive neuropsychological assessment. CMB number and topography (lobar, deep or infratentorial, or mixed location) were rated. WMHs were quantified with volumetric analysis.

**Results:** Compared to absence of CMBs, higher burden of CMBs (≥3) was associated independently of age with worse performance in processing speed (stand. β=0.18–0.23, p<0.05) and executive functions (stand. β=0.18–[–0.25], p<0.05), but not with episodic memory. Mild WMHs had no independent relationships with cognition. Compared to strictly lobar or deep or infratentorial CMBs, mixed location of CMBs was more often negatively related to cognitive performance (stand. β=0.20– 0.32, p<0.05).

**Conclusions:** CMBs were related to a subtle, yet systematic impairment in processing speed and executive functions, whereas no such association was observed for WMHs. The results provide insight into the development of early cSVD-related cognitive changes already in midlife and suggest an increased risk of cognitive decline in T1D.

## INTRODUCTION

Individuals with type 1 diabetes (T1D) have an elevated risk of microvascular complications, such as diabetic kidney disease, retinopathy, and cerebral small vessel disease (cSVD)^1,2^. CSVD affects brain’s microvessels and leads to diverse pathology seen on brain imaging, typically in the elderly. However, compared to healthy middle-aged adults, individuals with T1D more often manifest cSVD-related brain changes including white matter hyperintensities (WMHs)^3^, alterations in white matter microstructure^4^, reduced grey matter density^5^ and volume^6^, and cerebral microbleeds (CMBs) ^1,7^. We showed that 33% of middle-aged individuals with T1D had CMBs^7^, whereas in the healthy population of this age the CMB prevalence is approximately 5%^8^. T1D is also a recognized risk factor for clinically significant cognitive impairment in middle age, affecting most prominently fine motor and information processing speed and executive functions^9,10^. However, the relationship between different cSVD-related brain changes and specific cognitive impairments in T1D is not fully understood.

In middle-aged people with T1D, WMHs and white matter microstructural changes are shown to associate with deficits in fine motor and information processing speed, executive functions, working memory, and general cognitive functioning^3,4,11,12^. In addition, smaller grey matter volumes in basal ganglia and cortical areas have been linked with cognitive impairments^13,14^. Although adults with T1D have a higher incidence of CMBs compared to controls^1^, research investigating their association with cognitive functions is sparse^12^.

CMBs have been shown to predict cognitive decline and dementia in older general populations^15,16^, although contrary results have also been reported^17^. Cognitive deficits have been shown in domains of executive functions, attention, information processing speed, and memory ^e.g.18,19^. Evidence on associations between CMB location and cognitive performance in older individuals is mixed. While some studies have shown associations between strictly lobar, deep or infratentorial, or mixed locations and cognitive performance ^e.g.19–21^, not all have found deep or infratentorial^20^ or any CMB location^22^ to be associated with cognition.

We aimed to study, whether CMBs and WMHs, as most pronounced cSVD markers in our study sample^1,7^ are related to information processing speed, executive functions, and verbal memory in middle-aged individuals with T1D showing no overt neurological symptoms. We also examined whether strictly lobar, deep or infratentorial, and mixed locations of CMBs differently associate with cognition in these individuals.

## MATERIALS AND METHODS

### Participants and study protocol

This magnetic resonance imaging (MRI) study^1^ belongs to the longitudinal Finnish Diabetic Nephropathy (FinnDiane) Study (Supplemental Table 1). This study involves human participants and was approved by the Ethics Committee of Helsinki and Uusimaa Hospital District (HUS/3313/2018). The study was conducted in accordance with Declaration of Helsinki. Each participant signed a written informed consent.

The participants were initially recruited between 2011–2017. Primary inclusion criteria included age between 18 and 50 years, T1D onset before age of 40 years, and no end-stage kidney disease, history of overt neurological disease, or contraindications for brain MRI^1^ (Figure 1). All participants underwent a clinical examination, brain MRI^7^, and a comprehensive neuropsychological assessment in 2019–2024. The median time between brain MRI and neuropsychological assessment was 14.0 days (IQR 6.0–28.5), and the assessments occurred for 78% of the participants within a month and for 93% within three months.

**Figure 1.**
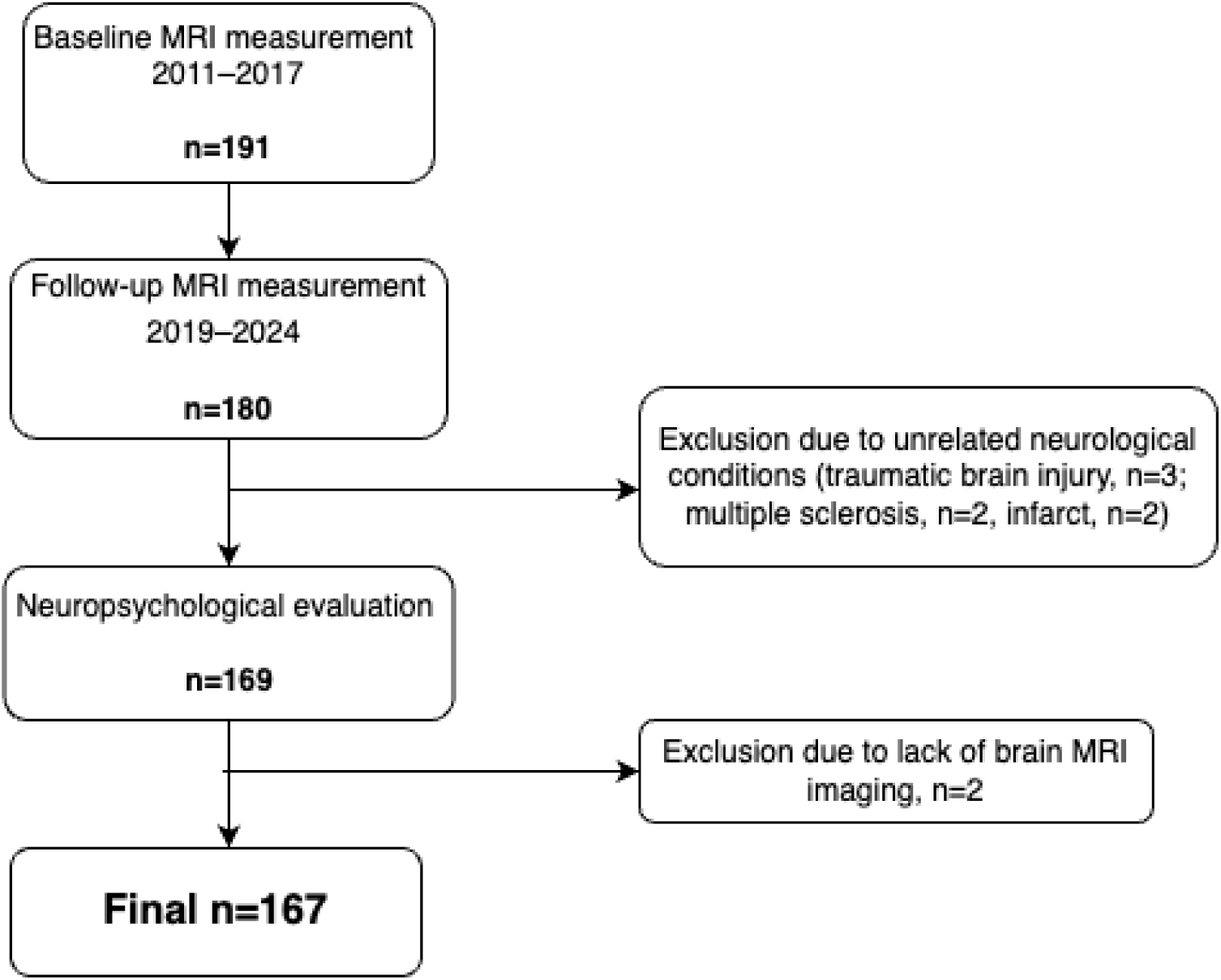
Flowchart of the participant recruitment and data acquisition for the current study.

### Clinical examinations

All participants attended clinical examination at the FinnDiane research unit at the Helsinki University Hospital^7^, covering evaluation of medical history, current medication, office blood pressure, waist circumference, and body mass index (Table 1 and Table 2). Glycated haemoglobin (HbA1c), low- and high-density lipoproteins, and creatinine were determined from blood samples. Estimated glomerular filtration rate was calculated with the Chronic Kidney Disease Epidemiology Collaboration (CKD-EPI) 2009 equation. Urinary albumin excretion rate was categorized based on two out of three timed overnight or 24-hour urine collections according to standard criteria. Furthermore, participants filled in questionnaires on demographic and lifestyle information^1^.

**Table 1.**
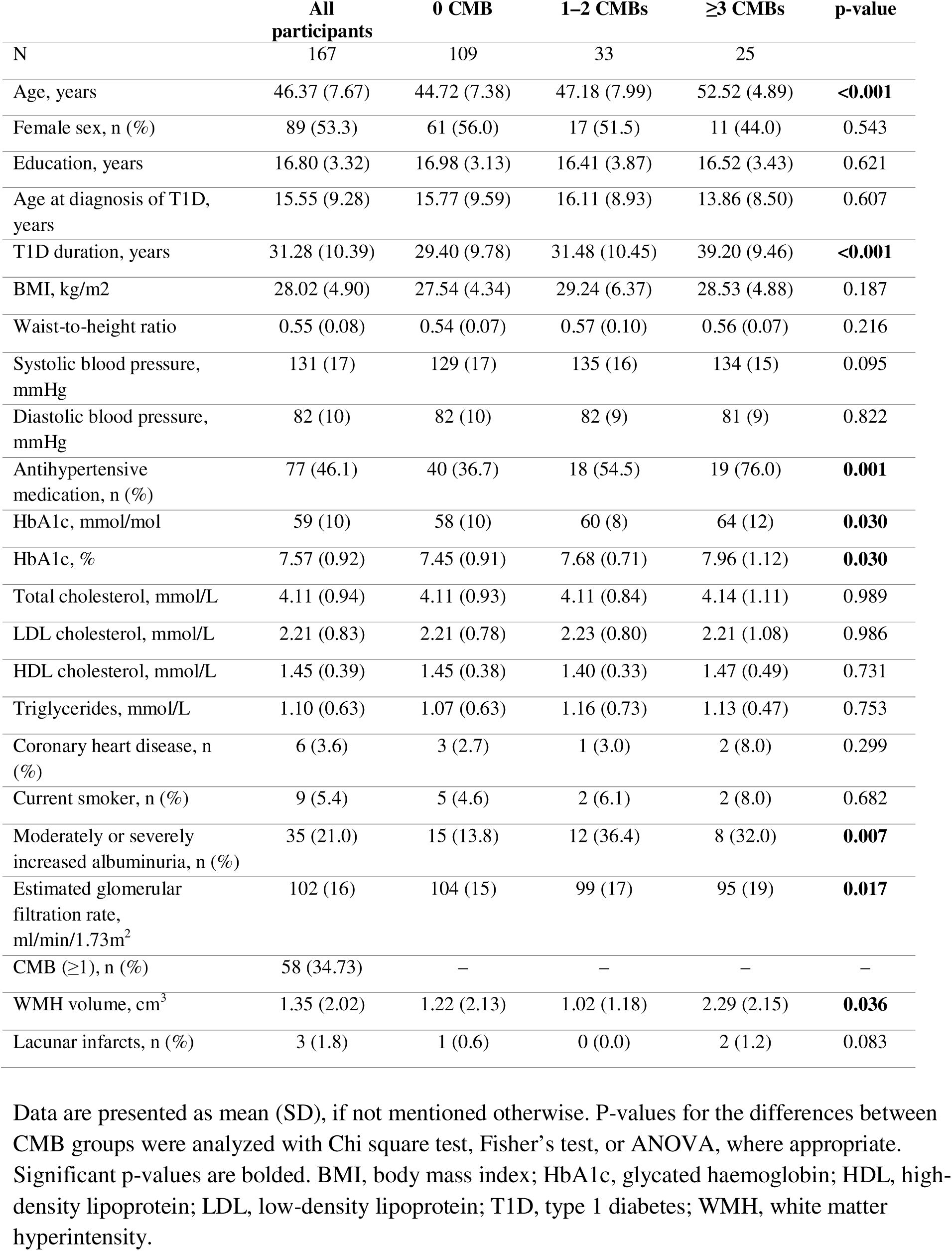
Demographic and clinical characteristics of all participants and participants in cerebral microbleed (CMB) groups at the clinical examination.

|  | All participants | 0 CMB | 1–2 CMBs | ≥3 CMBs | p-value |
| --- | --- | --- | --- | --- | --- |
| N | 167 | 109 | 33 | 25 |  |
| Age, years | 46.37 (7.67) | 44.72 (7.38) | 47.18 (7.99) | 52.52 (4.89) | **<0.001** |
| Female sex, n (%) | 89 (53.3) | 61 (56.0) | 17 (51.5) | 11 (44.0) | 0.543 |
| Education, years | 16.80 (3.32) | 16.98 (3.13) | 16.41 (3.87) | 16.52 (3.43) | 0.621 |
| Age at diagnosis of T1D, years | 15.55 (9.28) | 15.77 (9.59) | 16.11 (8.93) | 13.86 (8.50) | 0.607 |
| T1D duration, years | 31.28 (10.39) | 29.40 (9.78) | 31.48 (10.45) | 39.20 (9.46) | **<0.001** |
| BMI, kg/m<sup>2</sup> | 28.02 (4.90) | 27.54 (4.34) | 29.24 (6.37) | 28.53 (4.88) | 0.187 |
| Waist-to-height ratio | 0.55 (0.08) | 0.54 (0.07) | 0.57 (0.10) | 0.56 (0.07) | 0.216 |
| Systolic blood pressure, mmHg | 131 (17) | 129 (17) | 135 (16) | 134 (15) | 0.095 |
| Diastolic blood pressure, mmHg | 82 (10) | 82 (10) | 82 (9) | 81 (9) | 0.822 |
| Antihypertensive medication, n (%) | 77 (46.1) | 40 (36.7) | 18 (54.5) | 19 (76.0) | **0.001** |
| HbA1c, mmol/mol | 59 (10) | 58 (10) | 60 (8) | 64 (12) | **0.030** |
| HbA1c, % | 7.57 (0.92) | 7.45 (0.91) | 7.68 (0.71) | 7.96 (1.12) | **0.030** |
| Total cholesterol, mmol/L | 4.11 (0.94) | 4.11 (0.93) | 4.11 (0.84) | 4.14 (1.11) | 0.989 |
| LDL cholesterol, mmol/L | 2.21 (0.83) | 2.21 (0.78) | 2.23 (0.80) | 2.21 (1.08) | 0.986 |
| HDL cholesterol, mmol/L | 1.45 (0.39) | 1.45 (0.38) | 1.40 (0.33) | 1.47 (0.49) | 0.731 |
| Triglycerides, mmol/L | 1.10 (0.63) | 1.07 (0.63) | 1.16 (0.73) | 1.13 (0.47) | 0.753 |
| Coronary heart disease, n (%) | 6 (3.6) | 3 (2.7) | 1 (3.0) | 2 (8.0) | 0.299 |
| Current smoker, n (%) | 9 (5.4) | 5 (4.6) | 2 (6.1) | 2 (8.0) | 0.682 |
| Moderately or severely increased albuminuria, n (%) | 35 (21.0) | 15 (13.8) | 12 (36.4) | 8 (32.0) | **0.007** |
| Estimated glomerular filtration rate, ml/min/1.73m<sup>2</sup> | 102 (16) | 104 (15) | 99 (17) | 95 (19) | **0.017** |
| CMB (≥1), n (%) | 58 (34.73) | – | – | – | – |
| WMH volume, cm<sup>3</sup> | 1.35 (2.02) | 1.22 (2.13) | 1.02 (1.18) | 2.29 (2.15) | **0.036** |
| Lacunar infarcts, n (%) | 3 (1.8) | 1 (0.6) | 0 (0.0) | 2 (1.2) | 0.083 |

Data are presented as mean (SD), if not mentioned otherwise. P-values for the differences between CMB groups were analyzed with Chi square test, Fisher's test, or ANOVA, where appropriate. Significant p-values are bolded. BMI, body mass index; HbA1c, glycated haemoglobin; HDL, high-density lipoprotein; LDL, low-density lipoprotein; T1D, type 1 diabetes; WMH, white matter hyperintensity.

**Table 2.**
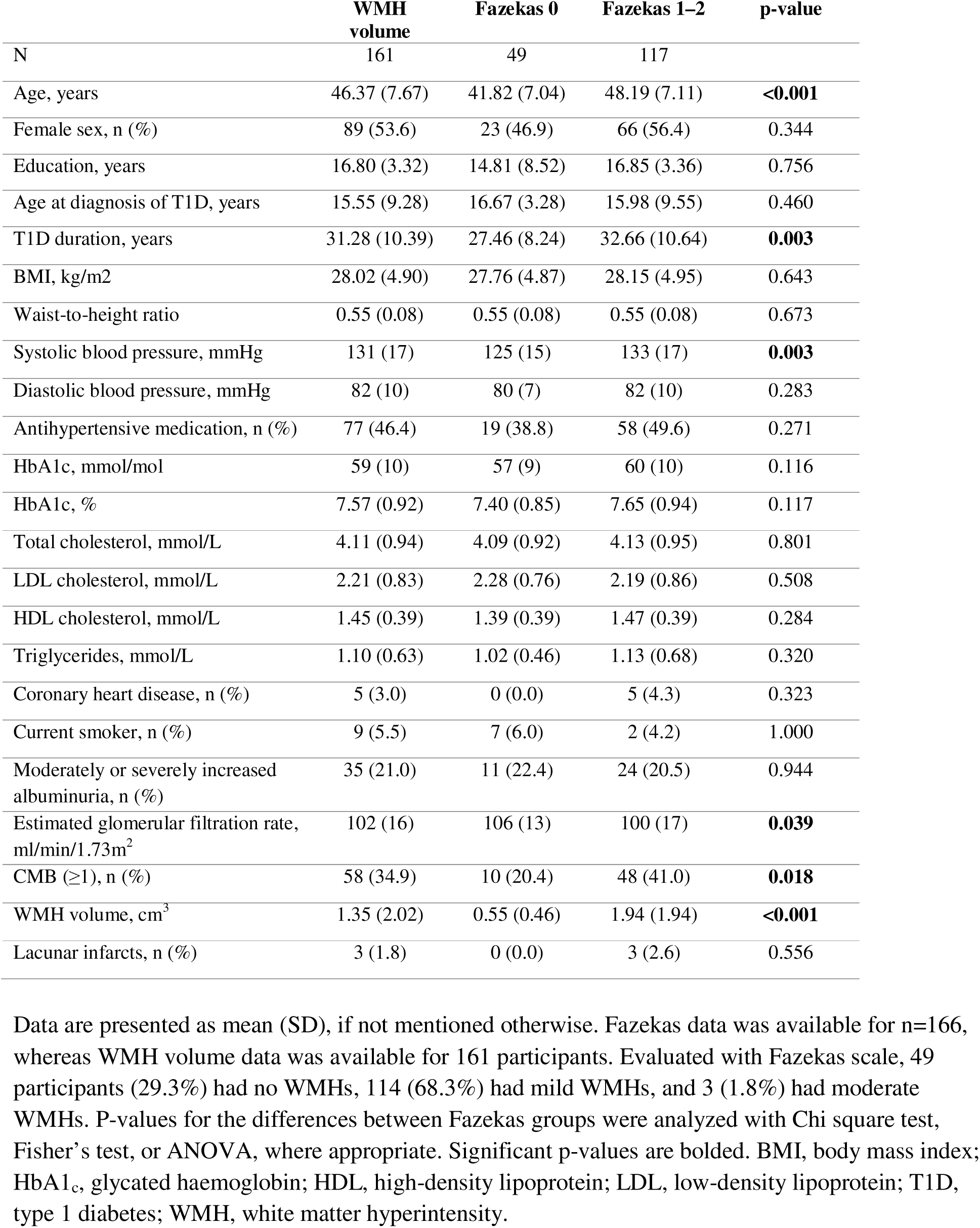
Demographic and clinical characteristics of all participants with available white matter hyperintensity (WMH) volumes and grouped according to WMH severity evaluated with Fazekas scale.

### Brain MRI

Data acquisition for the baseline CMB quantity data and CMB progression was performed during 2011–2017 at the Helsinki University Hospital’s Medical Imaging Center with 3T Achieva, Philips Ingenia (Netherlands)^1^. MRI sequences included T1, T2, FLAIR, susceptibility-weighted imaging, T2*, diffusion-weighted imaging, T1 Magnetization-Prepared Rapid Gradient Echo, and MR time-of-flight. The follow-up MRI was performed with a 3T Philips Ingenia (Best, The Netherlands) and a 32-channel head coil using the same sequences^7^.

The number and location of CMBs, as well as the severity of WMHs assessed with Fazekas scale and lacunes for descriptive purposes, were visually evaluated by a senior neuroradiologist (JM), who was blinded to clinical and neuropsychological evaluations, according to standard criteria^23,24^. In addition, WMHs were segmented from the 3D FLAIR with the Lesion prediction algorithm^25^ as implemented in the Lesion segmentation tool for SPM toolbox version 3.0.0 (www.statistical-modelling.de/lst.html). To correct for the brain size variation, we normalized WMH volumes by dividing them with the total intracranial volume^26^. The intracranial volume was obtained by segmenting the T1 3D and T2 3D TSE sequence Freesurfer (version 7.1.1) segmentation^27,28^. The segmentations were manually corrected by an experienced physicist (LK), if necessary.

### Neuropsychological assessment

Neuropsychological assessment (approximately two hours) was performed at follow-up and consisted of an extensive battery of established paper-and-pencil tests and computerized cognitive measures carried out by trained research psychologists. The present analysis focused on processing speed, executive functions, i.e., cognitive flexibility, inhibitory control, and working memory^29^, and verbal episodic memory. Cognitive tests are described in detail in Supplemental Table 2. Processing speed was evaluated with Wechsler Adult Intelligence Scale IV (WAIS-IV) Coding subtask^30^, Stroop Colour-Naming (Stroop-II)^31^, and Reaction Time (FAT-RT) and Numbers (FAT-N) subtests from the Flexible Attention Test^32^. FAT is a computerized tablet test including eight tasks of progressing difficulty covering the domains of visuomotor speed, attention, set shifting, and working memory, and has been shown to correlate logically with corresponding traditional neuropsychological tests^32,33^. Cognitive flexibility was assessed with FAT subtasks of Numbers and Letters (FAT-NL), Numbers and Shapes (FAT-NS), and Numbers and Months Forward (FAT-NMF) and Backward (FAT-NMB)^32^. Inhibitory control was measured with Stroop Colour-Incongruent (Stroop-III)^31^. Working memory was assessed with FAT Visuospatial Memory Span Forward (FAT-MF) and Backward (FAT-MB) as measures of visuospatial short-term/working memory^32,33^. Verbal episodic memory was measured with the Wechsler Memory Scale III (WMS-III) Word Lists of immediate (sum of four recalls) and delayed recall (WL-1 and WL-2, respectively)^34^. Participants were advised to avoid any hypoglycaemic events 24 hours prior to the assessment, and the blood glucose concentration had to be >3 mmol/L in the beginning of the assessment.

### Statistical analysis

Statistical analyses were performed with R version 4.2.1. Statistical significance was reported as p-value <0.05 and effect size with Cohen’s *f^2^* or *d*.

There were a few missing values in the cognitive test scores due to the participants’ inability to complete the task, or technical, or other reasons. The number of missing values ranged from 0 to 3, except for the FAT-MB, which had 17 missing values. Missing values in clinical descriptive data in Tables 1 and 2 ranged from 0 to 5 and were not replaced.

For descriptive analyses, Chi square test, Fisher’s test, t-test, Wilcoxon test, ANOVA, and Spearman(ρ) or Pearson(*r*) correlations were used where appropriate. We report the data as mean (standard deviation, sd), if not stated otherwise.

The main analyses were performed with linear regression models, which were all checked for their assumptions. Logarithmic transformations were performed for the dependent variables, where appropriate. Assumption of homoscedasticity was assessed visually, and no systematic evidence for violation was found. In the CMB location models, the prediction was more reliable for the groups of larger size (largest group size: n=109, smallest group size: n=11), which should be considered while interpreting the results. The residual normality assumption was slightly violated in some of the models, partly due to outliers. However, Cook’s value was <1 in all models. Removal of the influential data points removed the two significant associations in interaction models.

The main linear regression analyses between selected cSVD markers and cognition included cognitive test scores as dependent variables and separately CMBs and WMH volume as independent variables, with both models adjusted for age as the main confounder (Tables 1 and 2). Participants were classified into 3 groups by the number of CMBs (0, 1-2, ≥3)^16^, where 0 CMBs served as the reference category. WMH volume was treated as a continuous variable, and due to skewed distribution, logarithmic transformation was applied. We further performed sensitivity analyses to see whether the associations between cognition and CMBs were significant after controlling for main vascular and diabetes-related factors associated with CMBs (Tables 1 and 2). We performed three models adjusted for age and one by one estimated glomerular filtration rate (eGFR) ml/min/1.73m², systolic blood pressure, and HbA1_c_. Disease duration was not included into the analyses because of its strong correlation with age. Lacunes were not considered in the analyses due to their small incidence (n=3).

To further see the combined effect of selected brain cSVD markers, they were added as interaction terms (CMB group*WMH volume) into the multivariable model together with age as the main confounder.

To analyse the association between CMB location and cognition, we performed linear regression models with cognitive scores as dependent variables and CMB location as independent variable. CMB location was classified into four categories: no CMBs (n=109), strictly lobar (n=24), deep or infratentorial (n=11), and mixed (n=23) (categorized binarily: absent or present).

False discovery rate (FDR) correction was applied using the Benjamini-Hochberg procedure to control for multiple comparisons. Adjusted p-values of <0.05 were considered significant.

## RESULTS

### Participant characteristics

Of the initial 191 participants enrolled in the FinnDiane MRI substudy, 165 participants were available for analysis (Figure 1). Descriptive data of participants are presented in Tables 1 and 2. Forty-nine participants (29.3%) had none, 114 (68.3%) had mild, and three (1.8%) had moderate WMHs evaluated with Fazekas scale. WMH volumes were available from 161 participants (6 missing due to technical or data quality issues). Individuals with 17 missing values in FAT-MB did not differ from those with available data according to age, sex, years of education, absolute WMH volume, or CMBs (p>0.05).

### Associations between cognition, selected cSVD markers, and demographic variables

Demographic and vascular risk factors and cSVD brain changes for all the participants and for all the participants with available CMB data and in different CMB groups are described in Table 1. Similar information for all participants with available WMH volumes and for participants in different Fazekas groups are presented in Table 2.

Education was weakly associated with immediate verbal memory (WL-1 [ρ=0.27, p<0.001]), but not with other cognitive variables. Age at diabetes onset correlated weakly with delayed verbal memory (WL-2 [ρ=–0.19, p=0.013]). We also observed weak to moderate correlations between diabetes duration and processing speed (Coding [ρ=–0.26, p<0.001], FAT-R [ρ=0.38, p<0.001]), and FAT-N [ρ=0.26, p<0.001]), and executive functions with set shifting (FAT-NL [ρ=0.23, p=0.003], FAT-NS [ρ=0.23, p=0.003], FAT-NMF [ρ=0.29, p<0.001], and FAT-NMB [ρ=0.26, p=0.001]), inhibition (Stroop-III [ρ=0.15, p=0.049]), and visuospatial working memory (FAT-MB [ρ=–0.20, p=0.015]). HbA1c mm/mol correlated weakly with processing speed (Coding [*r*=–0.17, p=0.027] and FAT-R [ρ=0.20, p=0.010]) and executive function subdomain of set shifting (FAT-NMB [ρ=0.19, p=0.013]). Systolic blood pressure correlated moderately with episodic memory (WL-1 and WL-2 [ρ=–0.34 and –0.32, respectively, p<0.001 for both]) and weakly with processing speed (Coding [ρ=–0.19, p=0.014] and FAT-R [ρ=0.18, p=0.021]), and executive function subdomain of set shifting (FAT-NS [ρ=0.20, p=0.011] and FAT-NMB [ρ=0.20, p=0.011]). Diastolic blood pressure correlated weakly with delayed verbal memory (WL-2 [ρ=–0.16, p=0.043]), but not with other cognitive variables.

### Cross-sectional associations between selected cSVD brain markers and cognition

In multivariable analyses adjusting for age, 1–2 CMBs did not differ from 0 CMBs in any of the cognitive tests (Table 3). Instead, ≥3 CMBs were associated with processing speed (Coding, Stroop-II, and FAT-RT) and executive function subdomains of inhibitory control (Stroop-III), cognitive flexibility (FAT-NMB) and working memory (FAT-MB) (Model 1 in Table 3). The independent effect sizes for CMBs in these models were small (Cohen’s *f^2^*=0.04–0.06). After FDR correction (Supplemental Table 3), significant findings remained for processing speed (Stroop-II and FAT-RT), inhibitory control (Stroop-III) and working memory (FAT-MB). Those measures of processing speed (Coding) and executive functions (FAT-NMB), which did not show significant associations after FDR correction, demonstrated a near-significant trend (adjusted p-value=0.050). WMH volume was not associated with any of the cognitive measures (Model 2 in Table 3). The raw scores between CMB groups for tests with significant associations are presented in Figure 2.

**Figure 2.**
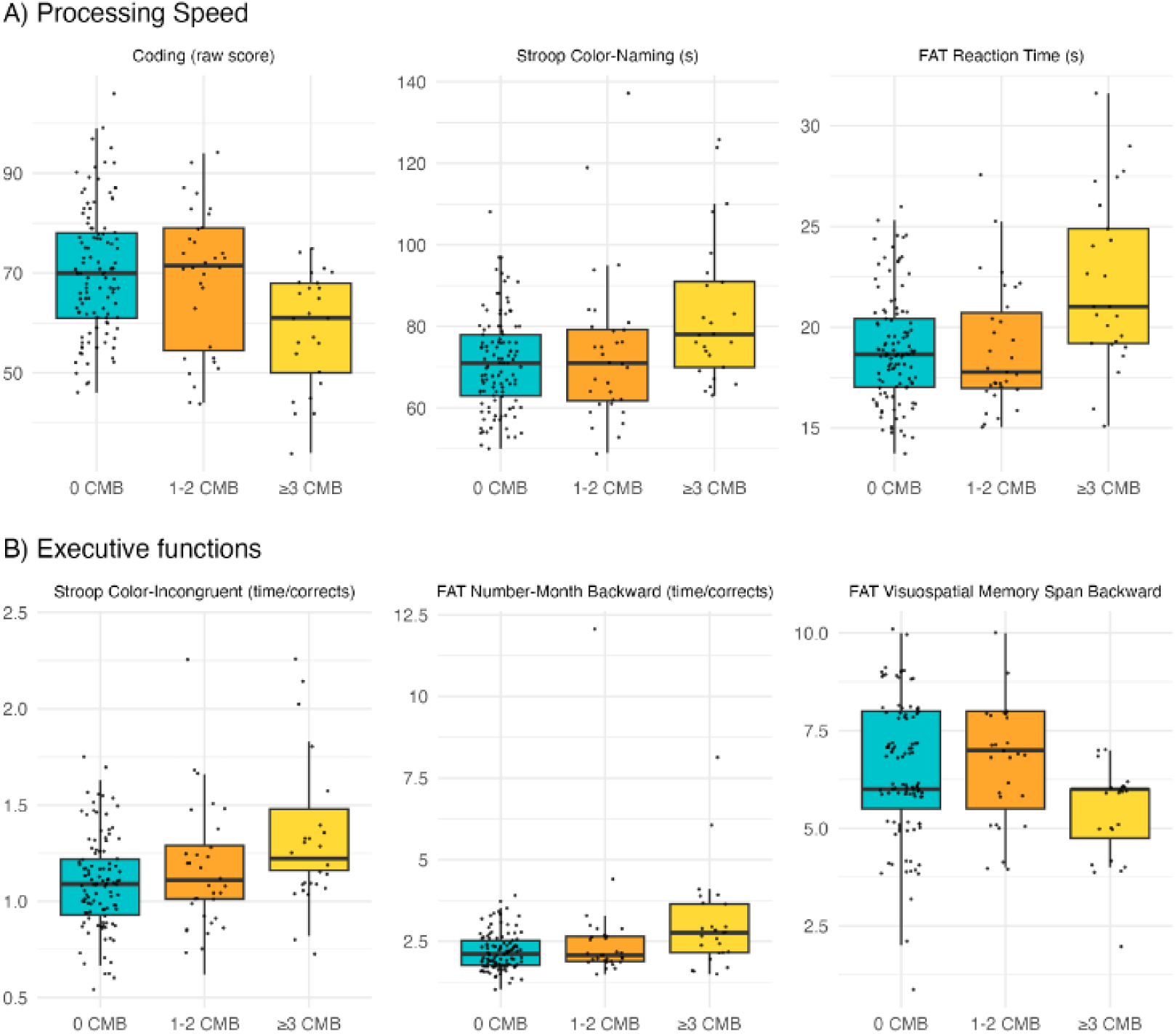
Performance in (A) processing speed and (B) executive function measures in groups based on number of cerebral microbleeds (CMB). In these tasks, ≥3 CMBs were associated with weaker performance compared to 0 CMBs, whereas 1–2 CMBs did not differ from 0 CMBs in terms of cognitive performance. FAT, Flexible Attention Test; FAT-MB, FAT Visuospatial Memory Span Backward; FAT-NMB, FAT Numbers and Months Backward; FAT-RT, FAT Reaction Time; Stroop-II, Stroop Colour-Naming; Stroop-III, Stroop Colour-Incongruent. Note that weaker performance is indicated in Coding and FAT Visuospatial Memory Span Backward as a smaller score, and in other tasks as a higher score, i.e., slower performance.

**Table 3.**
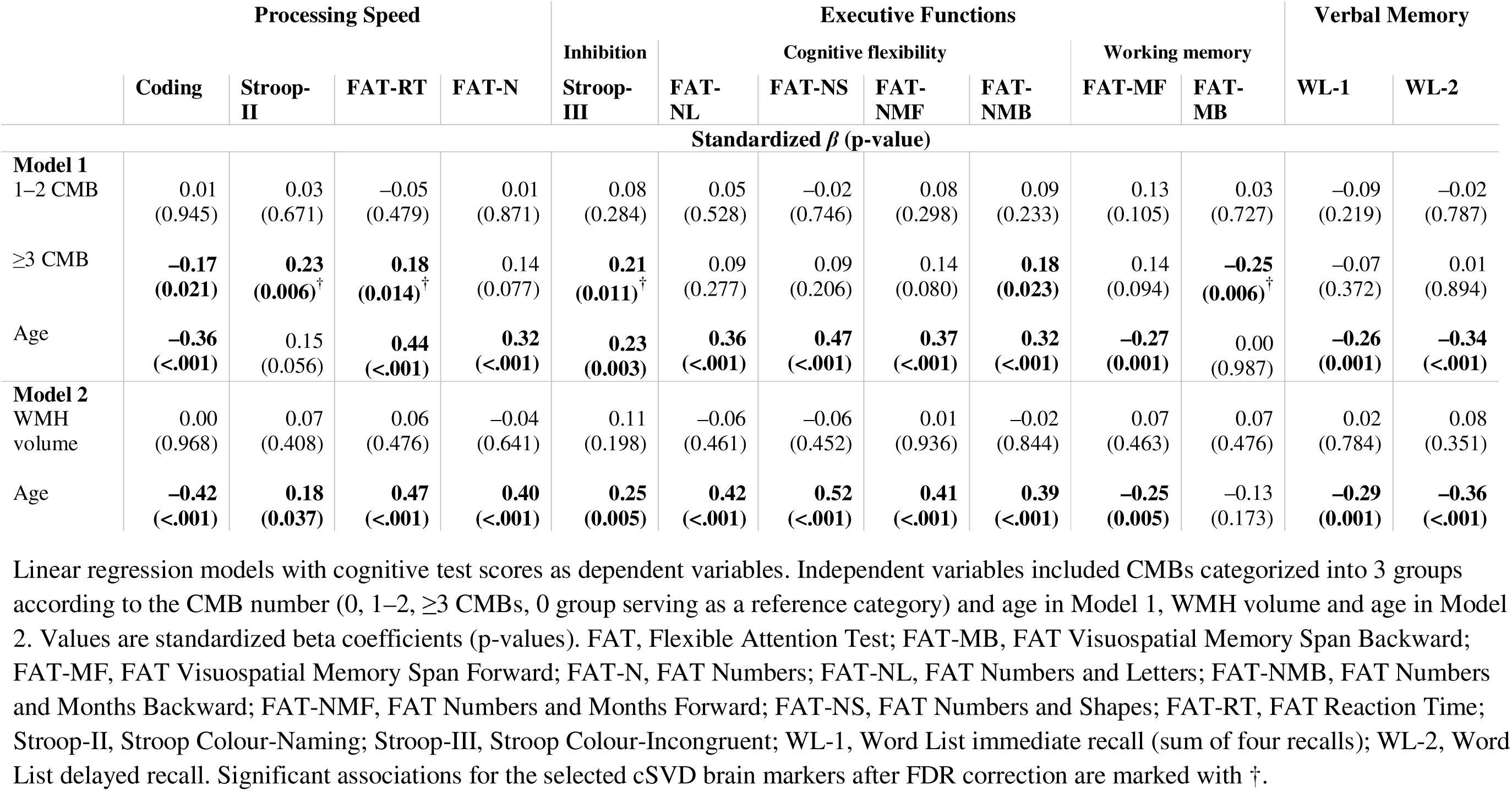
Cross-sectional associations of cerebral microbleeds (CMB) and white matter hyperintensity (WMH) volume with processing speed, executive functions, and verbal episodic memory, adjusted for age.

We further ran sensitivity analyses to assess the effect of confounding factors. Thus, Model 1 was further adjusted for eGFR, systolic blood pressure, and HbA1_c_, one covariate by one. After adjusting for differences among these clinical variables, those with ≥3 CMBs continued to have worse cognitive outcome in domains of processing speed and executive functions, compared to those without CMBs (Supplemental Tables 5, 6, and 7). After FDR correction, the associations mainly remained significant in the models adjusted for eGFR and systolic blood pressure, but not in the model adjusted for HbA1_c_.

### Combined effects of selected cSVD brain markers on cognition

In interaction models including cSVD markers as interaction terms (CMB group*WMH volume) we found that WMH volume moderated cognitive performance in processing speed (FAT-N; standardized β=–1.12, p=0.039) and executive flexibility (FAT-NS and FAT-NMB; standardized β=– 1.25, p=0.016 and standardized β=–1.24, p=0.023, respectively). The results indicated that in the group with ≥3 CMBs, those with larger WMH volume had poorer cognitive performance. However, the significant associations did not persist after FDR correction.

### Associations between CMB location and cognition

Mixed CMB location (n=23) was associated with multiple cognitive measures including processing speed (Coding, Stroop-II, FAT-RT, and FAT-N), executive function subcomponents of inhibitory control (Stroop-III), cognitive flexibility (FAT-NL, FAT-NS, FAT-NMF, and FAT-NMB), and working memory (FAT-MB), as well as immediate verbal memory (WL-1) (Table 5). Strictly lobar CMB location (n=24) was associated with processing speed (Coding), executive flexibility (FAT-NL, FAT-NMF, and FAT-NMB), and immediate verbal memory (WL-1). Deep or infratentorial location (n=11) was not associated with cognitive performance. The effect sizes for the significant models varied from small to close to moderate (Cohen’s *f^2^*=0.06–0.12). Cohen’s *d* effect size was calculated to compare the group with no CMBs with the other CMB location groups. For the mixed location, the significant results indicated moderate to large effect sizes (Cohen’s |*d*|=0.47–0.98). For the lobar location, the effect sizes varied from small to moderate-to-large (Cohen’s |*d*|=0.21–0.71). Further analyses revealed that single CMBs were seen in both lobar and deep or infratentorial locations, but ≥3 CMBs appeared most often in mixed locations (p<0.001, Supplemental Table 7). Most of the significant results (Table 4) remained after FDR correction.

**Table 4.** Associations of strictly lobar, deep or infratentorial, or mixed cerebral microbleed (CMB) location, with processing speed, executive functions, and verbal memory.

| Processing Speed |  |  |  |  | Executive Functions |  |  |  |  |  |  | Verbal Memory |  |
| --- | --- | --- | --- | --- | --- | --- | --- | --- | --- | --- | --- | --- | --- |
|  | Coding | Stroop-II | FAT-RT | FAT-N | Inhibition | Cognitive flexibility |  |  |  | Working memory |  | WL-1 | WL-2 |
|  |  |  |  |  | Stroop-III | FAT-NL | FAT-NS | FAT-NMF | FAT-NMB | FAT-MF | FAT-MB |  |  |
| Standardized $\beta$ (p-value) | | | | | | | | | | | | | |
| Strictly lobar | −0.07<br>(0.343) | 0.06<br>(0.417) | **0.17**<br>**(0.019)** | 0.07<br>(0.382) | 0.15<br>(0.056) | **0.16**<br>**(0.044)** | 0.08<br>(0.299) | **0.17**<br>**(0.027)** | **0.22**<br>**(0.005)<sup>†</sup>** | 0.04<br>(0.648) | −0.04<br>(0.646) | **−0.21**<br>**(0.006)<sup>†</sup>** | −0.15<br>(0.062) |
| Deep or infra-tentorial | 0.03<br>(0.729) | 0.01<br>(0.936) | −0.05<br>(0.527) | 0.00<br>(0.949) | 0.01<br>(0.874) | −0.03<br>(0.663) | 0.00<br>(0.984) | 0.00<br>(0.959) | 0.01<br>(0.946) | 0.13<br>(0.112) | 0.06<br>(0.439) | 0.06<br>(0.401) | 0.09<br>(0.274) |
| Mixed | **−0.32**<br>**($<0.001$)<sup>†</sup>** | **0.30**<br>**($<0.001$)<sup>†</sup>** | **0.32**<br>**($<0.001$)<sup>†</sup>** | **0.26**<br>**(0.001)<sup>†</sup>** | **0.28**<br>**($<0.001$)<sup>†</sup>** | **0.20**<br>**(0.012)<sup>†</sup>** | **0.23**<br>**(0.004)<sup>†</sup>** | **0.25**<br>**(0.001)<sup>†</sup>** | **0.24**<br>**(0.003)<sup>†</sup>** | 0.04<br>(0.591) | **−0.24**<br>**(0.003)<sup>†</sup>** | **−0.16**<br>**(0.040)** | −0.10<br>(0.190) |

Linear regression models with cognitive test scores as dependent variables. Independent variable, CMB location, was categorized into 4 categories of strictly lobar (n=24), deep or infratentorial (n=11), mixed (n=23), or no CMBs (n=109), which served as the reference group. FAT, Flexible Attention Test; FAT-MB, FAT Visuospatial Memory Span Backward; FAT-MF, FAT Visuospatial Memory Span Forward; FAT-N, FAT Numbers; FAT-NL, FAT Numbers and Letters; FAT-NMB, FAT Numbers and Months Backward; FAT-NMF, FAT Numbers and Months Forward; FAT-NS, FAT Numbers and Shapes; FAT-RT, FAT Reaction Time; Stroop-II, Stroop Colour-Naming; Stroop-III, Stroop Colour-Incongruent; WL-1, Word List immediate recall (sum of 4 recalls); WL-2, Word List delayed recall. Associations that remain significant after FDR correction are marked with <sup>†</sup>.

## DISCUSSION

This study assessed the associations between selected cSVD-related brain changes and cognitive performance among middle-aged adults with T1D. Our main finding was that in T1D, higher number of CMBs was associated with subtle, yet systematic deficits in processing speed and executive functions, but not in verbal episodic memory, independently of age and mild WMHs. Mixed location of CMBs was more prominently associated with cognitive performance compared to strictly lobar and deep or infratentorial locations. The presence of WMHs, which were of mild severity in most of the participants, were not associated with cognition when adjusting for age.

Those with ≥3 CMBs performed poorer in tasks of processing speed and executive functions, i.e., inhibition and working memory, but not in verbal episodic memory, compared to participants with no CMBs, when age as the main confounder was controlled. Cognitive performance did not differ between the group with 1–2 CMBs and the group with no CMBs. When we controlled for renal function and systolic blood pressure, the results remained significant. Nevertheless, adjusting for HbA1_c_ weakened the associations and they did not remain significant after FDR correction. However, since HbA1_c_ is associated with CMBs ^7^ and thus also accounts for a shared variance, this model may underestimate the association between CMBs and cognition. Earlier research on CMBs and cognition in individuals with T1D is mainly lacking. A single small-scale study found no association between the presence of baseline CMBs and general cognitive performance assessed 3.5 years later^12^. These differing results may be explained by our larger sample size, cross-sectional study design, domain-specific cognitive assessment, and quantification of CMBs. However, our results align with findings in an older general population, in which a higher number of CMBs was shown to be associated with poorer processing speed and executive functions, and more inconsistently with memory^18,19^.

Additionally, in older individuals, presence of CMBs has been shown to double the risk of cognitive decline and dementia^35^. The individuals with more CMBs did not perform poorer in verbal episodic memory, which is in line with earlier research in patients with T1D ^9,10^.

To our knowledge, earlier studies have not investigated the relations between CMB location and cognition in middle-aged individuals with T1D. In this study, we found that several CMBs were in both lobar and deep or infratentorial locations. Moreover, these numerous CMBs in the mixed location were widely associated with executive functions and processing speed, whereas single, strictly lobar CMBs were associated only with some cognitive tasks. Furthermore, deep or infratentorial CMBs were not linked with cognitive functioning. In older adults, not all earlier studies have found associations between deep or infratentorial, or any CMB location and cognition^20,22^, but negative associations between lobar, deep or infratentorial, and mixed CMB locations and cognitive functions have also been found ^e.g.19–21^. However, since most our individuals with mixed location had ≥3 CMBs, it remains unclear, whether the number of CMBs, CMB location, or both drive the cognitive deficits. Also, since the groups based on CMB location in our study were relatively small, our explorative results can only be interpreted indicatively and warrant more research.

Contrary to CMBs, WMH volume was not associated with cognition. This result differs from an earlier study in middle-aged individuals with T1D, which found associations between WMH severity and processing speed deficits^3^. Compared to that study, our participants had milder WMHs (mainly Fazekas grade 1), and they were not required to have childhood-onset T1D, which may partly explain the different findings. Among cognitively healthy young and middle-aged individuals, WMHs have not systematically been associated with cognition ^e.g.36,37^, but in older individuals, baseline WMH volume and WMH progression have been shown to predict cognitive decline and dementia^17,38^. We also found no evidence of a moderating effect of WMH volume on the relationship between CMBs and cognition.

There are some limitations to this study. First, cross-sectional setting limits us from drawing causal interpretations of the cSVD brain markers and cognitive functions. Second, our conclusions on working memory performance are limited to the visual domain. Additionally, the subgroups especially regarding the CMB location were small, and the results need to be replicated with larger sample sizes. Strengths of our study include the relatively large and well-defined sample, quantification of both CMBs and WMHs as the core cSVD lesion types with standard criteria, and the extensive neuropsychological test battery including computerized neuropsychological tests, enabling sensitive measurement of executive functions and processing speed. We performed multiple analyses due to the experimental nature of the studied phenomenon and because of this, we evaluated the effect sizes with the conventional Cohen’s measures and applied the Benjamini-Hochberg procedure to control for multiplicity.

In conclusion, our findings indicate that CMBs are associated with cognitive deficits in middle-aged individuals with T1D and suggest that the presence of multiple CMBs may serve as a marker of subtle cognitive deficits. In clinical practice, cognitive complaints in individuals with T1D manifesting cSVD-related brain changes should be taken into consideration. Interventional and follow-up studies are warranted to clarify how these early microvascular and cognitive symptoms may develop and could be prevented.

## Acknowledgements

We thank research psychologists Tuuli Levänen, Emma Talvitie, Enni Rasmus, and Heidi Heinonen for their expertise, and Anu Dufva, Anna Sandelin, and Kirsi Uljala for technical assistance. We are grateful to Pentti Pölönen and Oili Salonen for MRI work and sincerely thank all participants for making this study possible.

## Author contributions

IK, AT, LK, JM, LMT, JP, DG, JM, and HJ contributed to the study design, acquisition and interpretation of data. IK prepared first draft of the study. All authors reviewed and approved the final version of the manuscript.

## Funding

The FinnDiane Study was supported by grants from Folkhälsan Research Foundation, Wilhelm and Else Stockmann Foundation, Liv och Hälsa Society, Sigrid Jusélius Foundation (220027), Medical Society of Finland, and State funding for university-level health research by Helsinki University Hospital (TYH2023403). IK was supported by Kymenlaakso Regional Fund (500393 ERV323 and 500420 ERV224), the Diabetes Research Foundation, and Ministry of Social Affairs and Health/Kymenlaakso Central Hospital. DG was supported by Liv och Hälsa Society, Medical Society of Finland (Finska Läkaresällskapet), Sigrid Juselius Foundation, State Funding for University-level Health Research (TYH2021206), University of Helsinki, Minerva Foundation Institute for Medical Research, and Research Council of Finland (UAK1021MRI). HJ was funded by Research Council of Finland and Helsinki University Hospital (UAK2112JOK). None of the funding bodies had any role in the study design, collection, analysis, or interpretation of data, writing of the manuscript, or the decision to submit the manuscript for publication.

## Conflicts of Interest

IK, JM, LK, TIP, JV, LMT, and HJ report no competing interests. AT is a shareholder and co-founder of RokoteNyt Oy. P-HG has received investigator-initiated research grants from Eli Lilly and Roche, is an advisory board member for AbbVie, Astellas, AstraZeneca, Bayer, Boehringer Ingelheim, Cebix, Eli Lilly, Janssen, Medscape, Merck Sharp & Dohme, Mundipharma, Nestlé, Novartis, Novo Nordisk, and Sanofi; and has received lecture fees from Astellas, AstraZeneca, Bayer, Berlin Chemie, Boehringer Ingelheim, Eli Lilly, Elo Water, Genzyme, Merck Sharp&Dohme, Medscape, Menarini, Novartis, Novo Nordisk, PeerVoice, Sanofi, and Sciarc. TT is serving/has served as an advisory board member to AstraZeneca, Bayer, Bristol Myers Squibb, Boehringer Ingelheim, Inventiva, and Portola Pharma and received lecture honorarium from Argenx. JP reports Lecture or Advisory Board honoraria from Abbott, BMS-Pfizer, Bayer, Herantis Pharma, and Novo Nordisk. DG reports Lecture or Advisory Board Honoraria from Astellas, AstraZeneca, Bayer, Boehringer Ingelheim, Fresenius, GE Healthcare, Harald AI, Novo Nordisk, and Ratiopharm.

## Data availability statement

Individual-level data from the study participants are not publicly available due to consent restrictions provided by the participants at the time of data collection. Readers may propose collaboration to access the individual-level data by contacting the lead investigator.

## Notes

### Funding Statement

The FinnDiane Study was supported by grants from Folkhaelsan Research Foundation, Wilhelm and Else Stockmann Foundation, Liv och Haelsa Society, Sigrid Juselius Foundation (220027), Medical Society of Finland, and State funding for university-level health research by Helsinki University Hospital (TYH2023403). IK was supported by Kymenlaakso Regional Fund (500393 ERV323 and 500420 ERV224), the Diabetes Research Foundation, and Ministry of Social Affairs and Health/Kymenlaakso Central Hospital. DG was supported by Liv och Haelsa Society, Medical Society of Finland (Finska Laekaresaellskapet), Sigrid Juselius Foundation, State Funding for University-level Health Research (TYH2021206), University of Helsinki, Minerva Foundation Institute for Medical Research, and Research Council of Finland (UAK1021MRI). HJ was funded by Research Council of Finland and Helsinki University Hospital (UAK2112JOK). None of the funding bodies had any role in the study design, collection, analysis, or interpretation of data, writing of the manuscript, or the decision to submit the manuscript for publication.

### Author Declarations

This study involves human participants and was approved by the Ethics Committee of Helsinki and Uusimaa Hospital District (HUS/3313/2018).

