## Supplemental material for "CEREBRAL MICROBLEEDS ARE ASSOCIATED WITH DEFICITS IN COGNITIVE PROCESSING SPEED AND EXECUTIVE FUNCTIONS IN MIDDLE-AGED ADULTS WITH TYPE 1 DIABETES"

**SUPPLEMENTARY MATERIAL**

**Supplemental Table 1.** FinnDiane Study Centers and their physicians and nurses.

| FinnDiane Study Centers | Physicians and nurses |
|---|---|
| **Anjalankoski Health Center** | S.Koivula, T.Uggeldahl |
| **Central Finland Central Hospital, Jyväskylä** | T.Forslund, A.Halonen, A.Koistinen, P.Koskiaho, M.Laukkanen, J.Saltevo, M.Tiihonen |
| **Central Hospital of Åland Islands, Mariehamn** | M.Forsen, H.Granlund, A.-C.Jonsson, B.Nyroos |
| **Central Hospital of Kanta-Häme, Hämeenlinna** | P.Kinnunen, A.Orvola, T.Salonen, A.Vähänen |
| **Central Hospital of Kymenlaakso, Kotka** | R.Paldanius, M.Riihelä, L.Ryysy |
| **Central Hospital of Länsi-Pohja, Kemi** | H.Laukkanen, P.Nyländen, A.Sademies |
| **Central Ostrobothnian Hospital District, Kokkola** | S.Anderson, B.Asplund, U.Byskata, P.Liedes, M.Kuusela, T.Virkkala |
| **City of Espoo Health Center:** | |
| **Espoonlahti** | A.Nikkola, E.Ritola |
| **Tapiola** | M.Niska, H.Saarinen |
| **Samaria** | E.Oukko-Ruponen, T.Virtanen |
| **Viherlaakso** | A.Lyytinen |
| **City of Helsinki Health Center:** | |
| **Puistola** | H.Kari, T.Simonen |
| **Suutarila** | A.Kaprio, J.Kärkkäinen, B.Rantaeskola |
| **Töölö** | P.Kääriäinen, J.Haaga, A-L.Pietiläinen |
| **City of Hyvinkää Health Center** | S.Klemetti, T.Nyandoto, E.Rontu, S.Satuli-Autere |
| **City of Vantaa Health Center:** | |
| **Korso** | R.Toivonen, H.Virtanen |
| **Länsimäki** | R.Ahonen, M.Ivaska-Suomela, A.Jauhiainen |
| **Martinlaakso** | M.Laine, T.Pellonpää, R.Puranen |
| **Myyrmäki** | A.Airas, J.Laakso, K.Rautavaara |
| **Rekola** | M.Erola, E.Jatkola |
| **Tikkurila** | R.Lönnblad, A.Malm, J.Mäkelä, E.Rautamo |
| **Heinola Health Center** | P.Hentunen, J.Lagerstam |
| **Helsinki University Hospital, Department of Medicine, Division of Nephrology** | R.Bergdal, T.Claesson, A.Dufva, N.Elonen, M.Eriksson, J.Fagerudd, M.Feodoroff, D.Gordin, P.-H.Groop, O.Heikkilä, K.Hietala, S.Hägg-Holmberg, F.Jansson Sigfrids, M.Korolainen, J.Kytö, S.Lindh, J.Nicklén, H.Paajanen, K.Pettersson-Fernholm, K.Rimpeläinen, M.Rosengård-Bärlund, M.Rönnback, L.Salovaara, A.Sandelin, M.Saraheimo, S.Satuli-Autere, R.Simonsen, P.Smidtslund, L.Thorn, H.Tikkanen, J.Tuomikangas, A.Tynjälä, K.Uljala, T.Vesisenaho, J.Wadén, A.Ylinen |
| **Herttoniemi Hospital, Helsinki** | V.Sipilä |
| **Hospital of Lounais-Häme, Forssa** | T.Kalliomäki, J.Koskelainen, R.Nikkanen, N.Savolainen, H.Sulonen, E.Valtonen |
| **Hyvinkää Hospital** | L. Norvio, A.Hämäläinen |
| **Iisalmi Hospital** | E.Toivanen |
| **Jokilaakso Hospital, Jämsä** | A.Parta, I.Pirttiniemi |

| | |
|---|---|
| **Jorvi Hospital, Helsinki University Central Hospital** | S.Aranko, S.Ervasti, R.Kauppinen-Mäkelin, A.Kuusisto, T.Leppälä, K.Nikkilä, L.Pekkonen |
| **Jyväskylä Health Center, Kyllö** | K.Nuorva, M.Tiihonen |
| **Kainuu Central Hospital, Kajaani** | S.Jokelainen, K.Kananen, M.Karjalainen, P.Kemppainen, A-M.Mankinen, A.Reponen, M.Sankari |
| **Kerava Health Center** | H.Stuckey, P.Suominen |
| **Kirkkonummi Health Center** | A.Lappalainen, M.Liimatainen, J.Santaholma |
| **Kivelä Hospital, Helsinki** | A.Aimolahti, E.Huovinen |
| **Koskela Hospital, Helsinki** | V.Ilkka, M.Lehtimäki |
| **Kotka Health Center** | E.Pälikkö-Kontinen, A.Vanhanen |
| **Kouvola Health Center** | E.Koskinen, T.Siitonen |
| **Kuopio University Hospital** | E.Huttunen, R.Ikäheimo, P.Karhapää, P.Kekäläinen, M.Laakso, T.Lakka, E.Lampainen, L.Moilanen, S. Tanskanen, L.Niskanen, U.Tuovinen, I.Vauhkonen, E.Voutilainen |
| **Kuusamo Health Center** | T.Kääriäinen, E.Isopoussu |
| **Kuusankoski Hospital** | E.Kilkki, I.Koskinen, L.Riihelä |
| **Laakso Hospital, Helsinki** | T.Meriläinen, P.Poukka, R.Savolainen, N.Uhlenius |
| **Lahti City Hospital** | A.Mäkelä, M.Tanner |
| **Lapland Central Hospital, Rovaniemi** | L.Hyvärinen, K.Lampela, S.Pöykkö, T.Rompasaari, S.Severinkangas, T.Tulokas |
| **Lappeenranta Health Center** | P. Erola, L.Härkönen, P.Linkola, T.Pekkanen, I.Pulli, E.Repo |
| **Lohja Hospital** | T.Granlund, K.Hietanen, M.Porrassalmi, M.Saari, T.Salonen, M.Tiikkainen, |
| **Länsi-Uusimaa Hospital, Tammisaari** | I.-M.Jousmaa, J.Rinne |
| **Loimaa Health Center** | A.Mäkelä, P.Eloranta |
| **Malmi Hospital, Helsinki** | H.Lanki, S.Moilanen, M.Tilly-Kiesi |
| **Mikkeli Central Hospital** | A.Gynther, R.Manninen, P.Nironen, M.Salminen, T.Vänttinen |
| **Mänttä Regional Hospital** | I.Pirttiniemi, A-M.Hänninen |
| **North Karelian Hospital, Joensuu** | U-M.Henttula, P.Kekäläinen, M.Pietarinen, A.Rissanen, M.Voutilainen |
| **Nurmijärvi Health Center** | A.Burgos, K.Urtamo |
| **Oulaskangas Hospital, Oulainen** | E.Jokelainen, P-L.Jylkkä, E.Kaarlela, J.Vuolaspuro |
| **Oulu Health Center** | L.Hiltunen, R.Häkkinen, S.Keinänen-Kiukaanniemi |
| **Oulu University Hospital** | R.Ikäheimo |
| **Päijät-Häme Central Hospital** | H.Haapamäki, A.Helanterä, S.Hämäläinen, V.Ilvesmäki, H.Miettinen |
| **Palokka Health Center** | P.Sopanen, L.Welling |
| **Pieksämäki Hospital** | V.Sevtsenko, M.Tamminen |
| **Pietarsaari Hospital** | M-L.Holmbäck, B.Isomaa, L.Sarelin |
| **Pori City Hospital** | P.Ahonen, P.Merisalo, E.Muurinen, K.Sävelä |
| **Porvoo Hospital** | M.Kallio, B.Rask, S.Rämö |
| **Raahe Hospital** | A.Holma, M.Honkala, A.Tuomivaara, R.Vainionpää |
| **Rauma Hospital** | K.Laine, K.Saarinen, T.Salminen |
| **Riihimäki Hospital** | P.Aalto, E.Immonen, L.Juurinen |
| **Salo Hospital** | A.Alanko, J.Lapinleimu, P.Rautio, M.Virtanen |
| **Satakunta Central Hospital, Pori** | M.Asola, M.Juhola, P.Kunelius, M.-L.Lahdenmäki, P.Pääkkönen, M.Rautavirta |

| | |
|---|---|
| **Savonlinna Central Hospital** | T.Pulli, P.Sallinen, M.Taskinen, E.Tolvanen, T.Tuominen, H.Valtonen, A.Vartia, S-L.Viitanen |
| **Seinäjoki Central Hospital** | O.Antila, E.Korpi-Hyövälti, T.Latvala, E.Leijala, T.Leikkari, M.Punkari N.Rantamäki, H.Vähävuori |
| **South Karelia Central Hospital, Lappeenranta** | T.Ensala, E.Hussi, R.Härkönen, U.Nyholm, J.Toivanen |
| **Tampere Health Center** | A.Vaden, P.Alarotu, E.Kujansuu, H.Kirkkopelto-Jokinen, M.Helin, S.Gummerus, L.Calonius, T.Niskanen, T.Kaitala, T.Vatanen |
| **Tampere University Hospital** | P. Hannula, I.Ala-Houhala, R.Kannisto, T.Kuningas, P.Lampinen, M.Määttä,H.Oksala, T.Oksanen, A.Putila, H.Saha, K.Salonen, H.Tauriainen, S.Tulokas |
| **Tiirismaa Health Center, Hollola** | T.Kivelä, L.Petlin, L.Savolainen |
| **Turku Health Center** | A.Artukka, I.Hämäläinen, L.Lehtinen, E.Pyysalo, H.Virtamo, M.Viinikkala, M.Vähätalo |
| **Turku University Central Hospital** | K.Breitholz, R.Eskola, K.Metsärinne, U.Pietilä, P.Saarinen, R.Tuominen, S.Äyräpää |
| **Vaajakoski Health Center** | K.Mäkinen, P.Sopanen |
| **Valkeakoski Regional Hospital** | S.Ojanen, E.Valtonen, H.Ylönen, M.Rautiainen, T.Immonen |
| **Vammala Regional Hospital** | I.Isomäki, R.Kroneld, L.Mustaniemi, M.Tapiolinna-Mäkelä |
| **Vasa Central Hospital** | S.Bergkulla, U.Hautamäki, V-A.Myllyniemi, I.Rusk |

**Supplemental Table 2.** Neuropsychological test variables.

| | | Description of the test | Variables | Cognitive function |
|---|---|---|---|---|
| **WAIS-IV Coding**[1] | | A sequence of numbers, each paired with a corresponding hieroglyphic-like symbol. Using a key, the examinee writes the symbol corresponding to its number. | Number of correct responses in a 2-minute time limit | Processing speed |
| **Flexible Attention Test**[2,3] | | | | |
| 1 | Reaction Time | 24 light grey circles are randomly distributed on the touch screen. One circle at the time turns green. The task is to tap the green circle as quickly as possible. | Total time to complete the task (s) | Processing speed |
| 2 | Numbers | Numbers 1-24 are scattered in circles on the touch screen. The task is to tap the numbers in order from smallest to highest as quickly as possible. | Total time to complete the task (s) | Processing speed |
| 3 | Numbers and Letters | Numbers 1-12 and alphabets A-L are randomly distributed in circles on the touch screen. The task is to alternately tap numbers and letters in sequence of 1-A-2-B-3-C, etc. | Total time to complete the task (s) divided by number of correct responses (max. 24) | Executive functions (Cognitive flexibility) |
| 4 | Numbers and Shapes | Numbers 1-12 in circles and another set of numbers 1-12 in squares are randomly distributed on the touch screen. The task is to tap the numbers from smallest to largest, alternating between circles and squares. | Total time to complete the task (s) divided by number of correct responses (max. 12) | Executive functions (Cognitive flexibility) |
| 5 | Numbers and Months Forward | Numbers 1-12 and months Jan-Dec are randomly distributed in circles on the touch screen. The task is to alternately tap numbers and months in forward order in a sequence of 1-Jan-2-Feb-3-Mar, etc. | Total time to complete the task (s) divided by number of correct responses (max. 24) | Executive functions (Cognitive flexibility) |
| 6 | Numbers and Months Backward | Numbers 1-12 and months Jan-Dec are randomly distributed in circles on the touch screen. The task is to alternately tap numbers and months in backward order in a sequence of 1-Dec-2-Nov-3-Oct, etc. | Total time to complete the task (s) divided by number of correct responses (max. 24) | Executive functions (Cognitive flexibility) |
| 7 | Visuospatial Memory Span Forward | Corsi Block-Tapping type forward span task: 24 light grey circles are distributed on the screen, one at a time turning red. The task is to tap the same circles in the same order. The length of the sequence to be recalled gradually increases until the subject makes two consecutive errors in the same span length. | Maximum forward span score | Executive functions (Visuospatial working memory) |
| 8 | Visuospatial Memory Span Backward | Corsi Block-Tapping type backward span task: 24 light grey circles are distributed on the screen, one at a time turning red. The task is to tap the same circles, but in reverse order. The length of the sequence to be recalled gradually increases until the subject makes two consecutive errors in the same span length. | Maximum backward span score | Executive functions (Visuospatial working memory) |
| **Stroop Color-Naming**[4,5] | | 100 colours (written as XXXX) are printed in either red, green, blue, or yellow. The task is to name the colours as fast as possible. | Total time to complete the task (s) | Processing speed |
| **Stroop Color-Incongruent**[4,5] | | 100 coloured words are written so that no word for a colour matches the ink colour (e.g., the word "blue" printed in red ink). The task is to name the ink colour of the word as fast and precise as possible, while inhibiting the word meaning. | Total time to complete the task (s) divided by number of correct responses (max. 100) | Executive functions (Inhibition) |
| **WMS-III Word List of immediate recall**[6] | | A list of 12 unrelated words is orally presented and the examinee is asked to recall as many as possible. This process is repeated across 4 learning trials. A new 12-word list is introduced once using the same procedure. After this, the examinee is asked to recall as many words from the original list as they can. | Total score is the sum of words recalled across all 4 trials (max. 48) | Immediate verbal memory |
| **WMS-III Word List of delayed recall**[6] | | The examinee is asked to recall the first list learned in the immediate condition after a delay of 25-35 minutes. | Total score is the number of words recalled after delay (max. 12) | Delayed verbal memory |

**Supplemental Table 3.** Original and False Discovery Rate (FDR) adjusted p-values for selected cross-sectional results.

| | **Processing Speed** | | | | **Executive Functions** | | | | | | | **Verbal memory** | |
|---|---|---|---|---|---|---|---|---|---|---|---|---|---|
| | **Coding** | **Stroop -II** | **FAT-RT** | **FAT-N** | **Stroop -III** | **FAT-NL** | **FAT-NS** | **FAT-NMF** | **FAT-NMB** | **FAT-MF** | **FAT-MB** | **WL-1** | **WL-2** |
| **Model 1** | | | | | | | | | | | | | |
| Original p-value | **0.021** | **0.006** | **0.014** | 0.077 | **0.011** | 0.277 | 0.206 | 0.080 | **0.023** | 0.094 | **0.006** | 0.372 | 0.894 |
| Adjusted p-value | 0.050 | **0.039** | **0.046** | 0.130 | **0.046** | 0.327 | 0.268 | 0.130 | 0.050 | 0.1357 | **0.039** | 0.403 | 0.894 |

Original and FDR adjusted p-values for ≥3 CMBs in linear regression models with cognitive test scores as dependent variables. Independent variables included cerebral microbleeds (CMB; categorized into three groups: 0, 1-2, ≥3 CMBs) and age in Model 1. Results are shown for group with ≥3 CMBs only, as participants with 1-2 CMBs did not differ from the reference group (0 CMBs) in any of the cognitive measures. FAT, Flexible Attention Test; FAT-MB, FAT Visuospatial Memory Span Backward; FAT-MF, FAT Visuospatial Memory Span Forward; FAT-N, FAT Numbers; FAT-NL, FAT Numbers and Letters; FAT-NMB, FAT Numbers and Months Backward; FAT-NMF, FAT Numbers and Months Forward; FAT-NS, FAT Numbers and Shapes; FAT-RT, FAT Reaction Time; Stroop-II, Stroop Colour-Naming; Stroop-III, Stroop Colour-Incongruent; WL-1, Word List immediate recall (sum of 4 recalls); WL-2, Word List delayed recall.

**Supplemental Table 4.** Cross-sectional associations of cerebral microbleeds (CMB) with processing speed, executive functions, and verbal memory, adjusted for age and estimated glomerular filtration rate (eGFR).

| | **Processing Speed** | | | | **Executive Functions** | | | | | | | **Verbal Memory** | |
|---|---|---|---|---|---|---|---|---|---|---|---|---|---|
| | | | | | **Inhibition** | **Cognitive flexibility** | | | | **Working memory** | | | |
| | **Coding** | **Stroop-II** | **FAT-RT** | **FAT-N** | **Stroop-III** | **FAT-NL** | **FAT-NS** | **FAT-NMF** | **FAT-NMB** | **FAT-MF** | **FAT-MB** | **WL-1** | **WL-2** |
| | **Standardized $\beta$ (p-value)** | | | | | | | | | | | | |
| 1–2 CMB | 0.00 (0.947) | 0.04 (0.615) | -0.05 (0.473) | 0.01 (0.900) | 0.09 (0.260) | 0.05 (0.517) | -0.02 (0.784) | 0.08 (0.268) | 0.09 (0.249) | 0.13 (0.103) | 0.03 (0.717) | -0.09 (0.252) | -0.02 (0.793) |
| ≥3 CMB | **-0.18 (0.022)** | **0.24 (0.004)**† | **0.18 (0.015)**† | 0.14 (0.086) | **0.21 (0.009)**† | 0.09 (0.269) | 0.10 (0.185) | 0.14 (0.067) | **0.17 (0.027)** | 0.14 (0.091) | **-0.24 (0.007)**† | -0.07 (0.408) | 0.01 (0.891) |
| Age | **-0.37 (<0.001)** | **0.20 (0.021)** | **0.43 (<0.001)** | **0.29 (<0.001)** | **0.27 (0.002)** | **0.37 (<0.001)** | **0.50 (<0.001)** | **0.41 (<0.001)** | **0.29 (<0.001)** | **-0.26 (0.004)** | 0.05 (0.582) | **-0.22 (0.011)** | **-0.34 (0.001)** |
| eGFR, ml/min/1.73m² | 0.00 (0.953) | 0.12 (0.158) | -0.02 (0.812) | -0.06 (0.460) | 0.09 (0.293) | 0.03 (0.703) | 0.08 (0.319) | 0.10 (0.200) | -0.08 (0.343) | 0.03 (0.730) | 0.13 (0.125) | 0.09 (0.270) | 0.01 (0.939) |

Linear regression models with cognitive test scores as dependent variables. Independent variables included CMBs categorized into 3 groups according to the CMB number (0, 1–2, ≥3 CMBs, 0 group serving as a reference category), and age and eGFR ml/min/1.73m² as covariates. Values are standardized beta coefficients (p-values). FAT, Flexible Attention Test; FAT-MB, FAT Visuospatial Memory Span Backward; FAT-MF, FAT Visuospatial Memory Span Forward; FAT-N, FAT Numbers; FAT-NL, FAT Numbers and Letters; FAT-NMB, FAT Numbers and Months Backward; FAT-NMF, FAT Numbers and Months Forward; FAT-NS, FAT Numbers and Shapes; FAT-RT, FAT Reaction Time; Stroop-II, Stroop Colour-Naming; Stroop-III, Stroop Colour-Incongruent; WL-1, Word List immediate recall (sum of 4 recalls); WL-2, Word List delayed recall. Significant associations for ≥3 CMBs after FDR correction are marked with †.

**Supplemental Table 5.** Cross-sectional associations of cerebral microbleeds (CMB) with processing speed, executive functions, and verbal memory, adjusted for age and systolic blood pressure (SBP).

| | **Processing Speed** | | | | **Executive Functions** | | | | | | | **Verbal Memory** | |
|---|---|---|---|---|---|---|---|---|---|---|---|---|---|
| | | | | | **Inhibition** | **Cognitive flexibility** | | | | **Working memory** | | | |
| | **Coding** | **Stroop-II** | **FAT-RT** | **FAT-N** | **Stroop-III** | **FAT-NL** | **FAT-NS** | **FAT-NMF** | **FAT-NMB** | **FAT-MF** | **FAT-MB** | **WL-1** | **WL-2** |
| | **Standardized $\beta$ (p-value)** | | | | | | | | | | | | |
| 1–2 CMB | 0.01 (0.937) | 0.04 (0.623) | -0.05 (0.507) | 0.01 (0.871) | 0.09 (0.264) | 0.05 (0.475) | -0.03 (0.711) | 0.08 (0.276) | 0.08 (0.253) | 0.12 (0.135) | 0.02 (0.776) | -0.08 (0.321) | 0.00 (0.962) |
| ≥3 CMB | **-0.18 (0.022)**† | **0.23 (0.005)**† | **0.18 (0.015)**† | 0.14 (0.078) | **0.20 (0.012)**† | 0.08 (0.295) | 0.10 (0.201) | 0.13 (0.086) | **0.18 (0.022)**† | 0.14 (0.085) | **-0.25 (0.006)**† | -0.08 (0.311) | 0.00 (0.969) |
| Age | **-0.36 (<0.001)** | **0.18 (0.043)** | **0.45 (<0.001)** | **0.32 (<0.001)** | **0.25 (0.004)** | **0.39 (<0.001)** | **0.45 (<0.001)** | **0.39 (<0.001)** | **0.30 (<0.001)** | **-0.31 (<0.001)** | -0.02 (0.805) | **-0.17 (0.045)** | **-0.26 (0.003)** |
| SBP | -0.01 (0.920) | -0.06 (0.489) | -0.03 (0.682) | 0.00 (0.995) | -0.04 (0.599) | -0.07 (0.376) | 0.04 (0.626) | -0.04 (0.569) | 0.04 (0.585) | 0.09 (0.271) | 0.05 (0.544) | **-0.21 (0.012)** | **-0.18 (0.025)** |

Linear regression models with cognitive test scores as dependent variables. Independent variables included CMBs categorized into 3 groups according to the CMB number (0, 1–2, ≥3 CMBs, 0 group serving as a reference category), and age and SBP as covariates. Values are standardized beta coefficients (p-values). FAT, Flexible Attention Test; FAT-MB, FAT Visuospatial Memory Span Backward; FAT-MF, FAT Visuospatial Memory Span Forward; FAT-N, FAT Numbers; FAT-NL, FAT Numbers and Letters; FAT-NMB, FAT Numbers and Months Backward; FAT-NMF, FAT Numbers and Months Forward; FAT-NS, FAT Numbers and Shapes; FAT-RT, FAT Reaction Time; Stroop-II, Stroop Colour-Naming; Stroop-III, Stroop Colour-Incongruent; WL-1, Word List immediate recall (sum of 4 recalls); WL-2, Word List delayed recall. Significant associations for ≥3 CMBs after FDR correction are marked with †.

**Supplemental Table 6.** Cross-sectional associations of cerebral microbleeds (CMB) with processing speed, executive functions, and verbal memory, adjusted for age and glycated haemoglobin ($HbA1_c$).

| | Processing Speed | | | | Executive Functions | | | | | | | Verbal Memory | |
|---|---|---|---|---|---|---|---|---|---|---|---|---|---|
| | | | | | Inhibition | Cognitive flexibility | | | | Working memory | | | |
| | **Coding** | **Stroop-II** | **FAT-RT** | **FAT-N** | **Stroop-III** | **FAT-NL** | **FAT-NS** | **FAT-NMF** | **FAT-NMB** | **FAT-MF** | **FAT-MB** | **WL-1** | **WL-2** |
| | **Standardized $\beta$ (p-value)** | | | | | | | | | | | | |
| 1–2 CMB | 0.01 (0.859) | 0.03 (0.731) | -0.06 (0.354) | 0.01 (0.938) | 0.08 (0.286) | 0.05 (0.531) | -0.03 (0.681) | 0.07 (0.354) | 0.08 (0.295) | 0.14 (0.085) | 0.03 (0.692) | -0.10 (0.188) | -0.01 (0.856) |
| ≥3 CMB | **-0.16 (0.038)** | **0.22 (0.008)** | **0.15 (0.038)** | 0.13 (0.111) | **0.21 (0.012)** | 0.09 (0.286) | 0.08 (0.277) | 0.12 (0.128) | **0.16 (0.046)** | 0.16 (0.062) | **-0.23 (0.010)** | -0.09 (0.295) | 0.02 (0.774) |
| Age | **-0.36 (<0.001)** | 0.15 (0.066) | **0.42 (<0.001)** | **0.31 (<0.001)** | **0.23 (0.003)** | **0.36 (<0.001)** | **0.46 (<0.001)** | **0.36 (<0.001)** | **0.31 (<0.001)** | **-0.26 (0.002)** | 0.01 (0.946) | **-0.27 (<0.001)** | **-0.33 (<0.001)** |
| $HbA1_c$ | -0.09 (0.198) | 0.07 (0.346) | **0.17 (0.015)** | 0.07 (0.326) | 0.00 (0.981) | 0.00 (0.993) | 0.07 (0.318) | 0.10 (0.182) | 0.12 (0.098) | -0.10 (0.193) | -0.07 (0.365) | 0.08 (0.297) | -0.07 (0.325) |

Linear regression models with cognitive test scores as dependent variables. Independent variables included CMBs categorized into 3 groups according to the CMB number (0, 1–2, ≥3 CMBs, 0 group serving as a reference category), and age and $HbA1_c$ as covariates. Values are standardized beta coefficients (p-values). FAT, Flexible Attention Test; FAT-MB, FAT Visuospatial Memory Span Backward; FAT-MF, FAT Visuospatial Memory Span Forward; FAT-N, FAT Numbers; FAT-NL, FAT Numbers and Letters; FAT-NMB, FAT Numbers and Months Backward; FAT-NMF, FAT Numbers and Months Forward; FAT-NS, FAT Numbers and Shapes; FAT-RT, FAT Reaction Time; Stroop-II, Stroop Colour-Naming; Stroop-III, Stroop Colour-Incongruent; WL-1, Word List immediate recall (sum of 4 recalls); WL-2, Word List delayed recall. Significant associations for ≥3 CMBs did not remain significant after FDR correction.

**Supplemental Table 7.** Contingency table of the cerebral microbleed (CMB) group and CMB location variables.

| | No CMBs (n) | Deep or infratentorial CMBs (n) | Strictly lobar CMBs (n) | Mixed CMBs (n) | Total (n) |
|---|---|---|---|---|---|
| 0 CMB (n) | 109 | 0 | 0 | 0 | 109 |
| 1–2 CMB (n) | 0 | 11 | 21 | 1 | 33 |
| ≥3 CMB (n) | 0 | 0 | 3 | 22 | 25 |
| Total (n) | 109 | 11 | 24 | 23 | |

Contingency table showing how the CMBs are distributed in the brain (deep or infratentorial, strictly lobar, or mixed location) among different CMB groups (0, 1–2, ≥3 CMBs).
